# Personalized Input-Output Hidden Markov Models for Disease Progression Modeling

**DOI:** 10.1101/2020.07.17.20153510

**Authors:** Kristen A. Severson, Lana M. Chahine, Luba Smolensky, Kenney Ng, Jianying Hu, Soumya Ghosh

## Abstract

Disease progression models are important computational tools in healthcare and are used for tasks such as improving disease understanding, informing drug discovery, and aiding in patient management. Although many algorithms for time series modeling exist, healthcare applications face particular challenges such as small datasets, medication effects, disease heterogeneity, and a desire for personalized predictions. In this work, we present a disease progression model that addresses these needs by proposing a probabilistic time-series model that captures individualized disease states, personalized medication effects, disease-state medication effects, or any combination thereof. The model builds on the framework of an input-output hidden Markov model where the parameters are learned using a structured variational approximation. To demonstrate the utility of the algorithm, we apply it to both synthetic and real-world datasets. In the synthetic case, we demonstrate the benefits afforded by the proposed model as compared to standard techniques. In the real-world cases, we use two Parkinson’s disease datasets to show improved predictive performance when ground truth is available and clinically relevant insights that are not revealed via classic Markov models when ground truth is not available.

## 1 Introduction

Disease progression models are an important tool for understanding the characteristics and progression trends of a wide variety of diseases. Applications of disease progression models include drug development [1], patient care management [2], and better understanding disease mechanisms [3]. Prediction tasks, including disease progression as well as the occurrence of specific outcomes, is a critical element of all of the latter. Disease progression modeling faces challenges that lead to specific algorithmic requirements. Here, we highlight and discuss four specific challenges: small datasets, a modeling goal of uncovering states, confounding of symptom presentation due to medication effects, and a desire to personalize prediction. We discuss each of these in more detail below.

- ***Small datasets***. Often, the amount of data available for modeling tasks is limited due to various causes such as the cost of acquiring the data, the enrollment burden on patients, study attrition, and data privacy issues. These small datasets motivate the incorporation of prior knowledge into the model structure.
- ***Uncovering disease states***. In many scenarios, disease progression models aim to both learn a quantitative model of future patient state along with salient disease states. Although some diseases have clearly definable pathologic stages, such as in cancer, many diseases do not have this characterization and discovering such stages is an important research goal. These states can be used for downstream tasks such as cohort generation and improved disease understanding.
- ***Confounding due to medication effects***. The datasets used for disease progression modeling are often complicated by medication effects, particularly in settings leveraging observational data. Medications may alter the symptoms of the patient, the underlying disease state, or both.
- ***Personalized predictions***. For heterogeneous diseases, there are many factors which may impact a particular patient’s disease progression, necessitating the modeling of personalized trajectories.

Disease progression models that address the aforementioned critical factors are particularly relevant for chronic diseases which are heterogeneous in both their manifestations and progression. In this setting, simple predictors of future disease state are ineffective. Parkinson’s disease (PD) is a useful case study for understanding the importance of disease progression modeling. PD is a chronic progressive neurodegenerative disorder with heterogeneous symptoms that may effect both motor and non-motor function. Two clinical subtypes of PD have been proposed [4], however the stability and progression of these subtypes is not well understood [5]. There are no medications that cure PD, however medications that improve quality of life by addressing common symptoms are available. The degree of response to medications, duration of response, and tolerance of medications vary from patient to patient. Disease progression models for PD would enable all of the benefits discussed above: improved disease understanding, patient care management, and drug development. PD is characteristic of many chronic conditions (e.g. Alzheimer’s disease, diabetes, ALS) where no disease-modifying therapeutics are available and is used to demonstrate our proposed approach.

The goal of the work is to model disease progression while taking medication effects and personalized disease trajectories into account. Motivated by PD and many other conditions for which there are currently no disease-modifying therapeutics, we focus on the scenario where medication does not alter the underlying disease but does alter the observations.

### Generalizable Insights about Machine Learning in the Context of Healthcare

In this work, we propose a probabilistic disease progression modeling framework to address the needs discussed above. Our models have the ability to account for personalized state and medication effects while learning disease states. This enables us to model heterogeneity in the disease population, a common characteristic of complex diseases. By carefully choosing the model structure, the parameters of these models can be learned without access to large datasets. Probabilistic models are particularly useful for disease progression modeling when one of the goals is to learn underlying disease states. Probabilistic models also have the ability to incorporate prior information, provide natural handling for missing data, and enable parameter uncertainty calculations. Because of these advantages, our research focuses on adapting probabilistic time series models for disease progression modeling. We rely on black-box variational inference and employ variational approximations that retain the model structure to perform accurate learning and inference. We demonstrate our approach on a synthetic dataset as well as two Parkinson’s disease datasets.

## 2 A personalized and medication aware disease progression model

Our proposed model builds on the hidden Markov model (HMM). A hidden Markov model describes sequential data through a series of transitions between latent states, with each state describing distinct characteristics of the observed data instances. An HMM has two primary components: (1) the transition model which describes the evolution of states over time and (2) the observation model which describes the manifestation of the state in observed space. Here, the latent states are denoted by *z*, with *z*_*i*_ corresponding to patient *i*’s full trajectory and *z*_*i,t*_ corresponding to an element in that trajectory at time *t*. Analogously, observations are denoted by *x* and medications are denoted by *d*. See Table 1 for additional details of the mathematical notation.

**Table 1:** Summary of notation used throughout the paper.

|  |  |  |  |
| --- | --- | --- | --- |
| $N$ | Number of patients, indexed $i \in \{1, \dots, N\}$ | $\lambda$ | Variational parameters |
| $T$ | Number of time points, indexed $t \in \{1, \dots, T\}$ | $\theta$ | Model parameters |
| $D$ | Dimensionality of the observations per time point | $\pi$ | Initial state distribution |
| $K$ | Number of latent states | $A$ | State transition distributions |
| $x$ | Observed clinical data | $v_k$ | State medication effects |
| $z$ | Latent states | $\phi_k$ | State parameters including $\mu_k$, state means, and $\Sigma_k$, state covariance |
| $d$ | Observed medication data | | |
| $m$ | Personalized medication effects | | |
| $r$ | Personalized state effects | | |

In our work, we use a standard transition model,

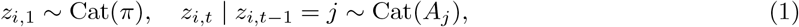

where 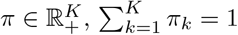 and 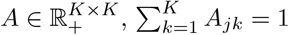. Cat(·) indicates a categorical distribution. An HMM with a Gaussian observation model is specified as *x*_*i,t*_ | *z*_*i,t*_ = *k* ∼ 𝒩 (*µ*_*k*_, Σ_*k*_), where 𝒩 (*µ*, Σ) represents a multivariate Gaussian distribution with mean *µ* ∈ ℝ^*D*^ and covariance Σ ∈ ℝ^*D×D*^. Below, we present observation models to account for systematic personalized and medication effects.

### Medication-aware HMMs

Assuming that medication information (*d*_*i,t*_) is available at each time step along with the observed data *x*_*i,t*_, the proposed observation model is

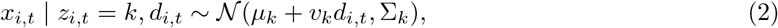

where *v*_*k*_ ∈ ℝ^*D×M*^ where *M* is the number of medications. The additive structure models state-specific deviations in the observed data from the disease state based on the presence of medication. The exact details of the implementation will depend on the expected impact of the drug. For instance, if the medication effect is not a function of the medication dose, *d*_*i,t*_ can be binary. Alternatively, the effect is scaled based on the dose. If the effect of medication does not vary along the disease trajectory, the model can be altered such that *v*_*k*_ = *v* ∀*k*.

For complex diseases, we expect medication responses to vary both among patients and also within the same patient along with the severity of the disease. We can modify Equation 2 to account for such effects by introducing state-specific effects for each patient, *x*_*i,t*_ | *z*_*i,t*_ = *k, d*_*i,t*_ ∼ 𝒩 (*µ*_*k*_ + *v*_*i,k*_*d*_*i,t*_, Σ_*k*_). However, the large number of parameters *v*_*i,k*_, makes learning challenging in such a model. Indeed, we expect to observe most patients in only a subset of all possible states, and inferences of *v*_*i,k*_ based on such data are likely to be unreliable. To circumvent this problem, we propose an alternate model where we factorize the medication effects into state-specific and patient-specific components,

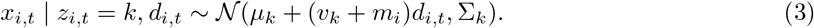

The state specific coefficients *v*_*k*_ are shared across observations assigned to state *k*, while *m*_*i*_ is shared across all observations from patient *i*. Such a factorization allows us to share statistical strength both between observations assigned to a common state and observations belonging to the same patient, thus providing more reliable inferences of personalized state specific medication effects.

### Personalized HMMs

To allow patients to deviate from the population at large, not as a function of medication, we introduce patient specific latent variables *r*_*i*_ ∈ ℝ^*D*^ to modify the mean response of patient *i*,

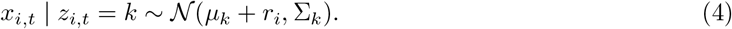

This enables personalization in how states might manifest in an individual.

It is natural to consider a more general variant that combines Equations 2 and 4 to account for both medication independent heterogeneity in the population as well as personalized medication effects,

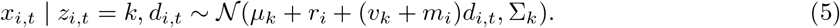

For ease of exposition, we frame the following discussion and the description of our learning algorithm in the context of this model (Equation 5) which subsumes the medication-aware and personalized models. Learning and inference in the models described via Equations 2 and 4 are analogous.

### Priors

We place Gaussian priors on the personalized effects (*m*_*i*_ and *r*_*i*_) and the state-specific medication effects (*v*_*k*_)

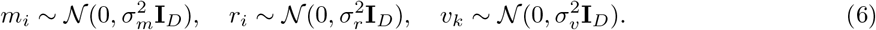

By employing zero mean Gaussian priors with appropriately chosen variances 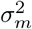 and 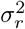 we encode our prior belief that while heterogeneity among patients and between states exists, the scale of this heterogeneity is small and the personalized effects do not deviate too far from the overall population. The prior over *v*_*k*_ regularizes its value and encourages the model to use the personalized effects *m*_*i*_ to explain the observed data.

Equations 1, 5, and 6 completely describe our model. The model parameters are 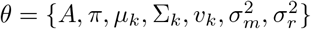 and the random variables are {*x, z, m, r*}. The resulting graphical model is shown in Figure 1. We refer to it as the personalized input-output hidden Markov model (PIOHMM) owing to its similarity to the input output HMM (IOHMM) [6]^1^.

**Figure 1:**
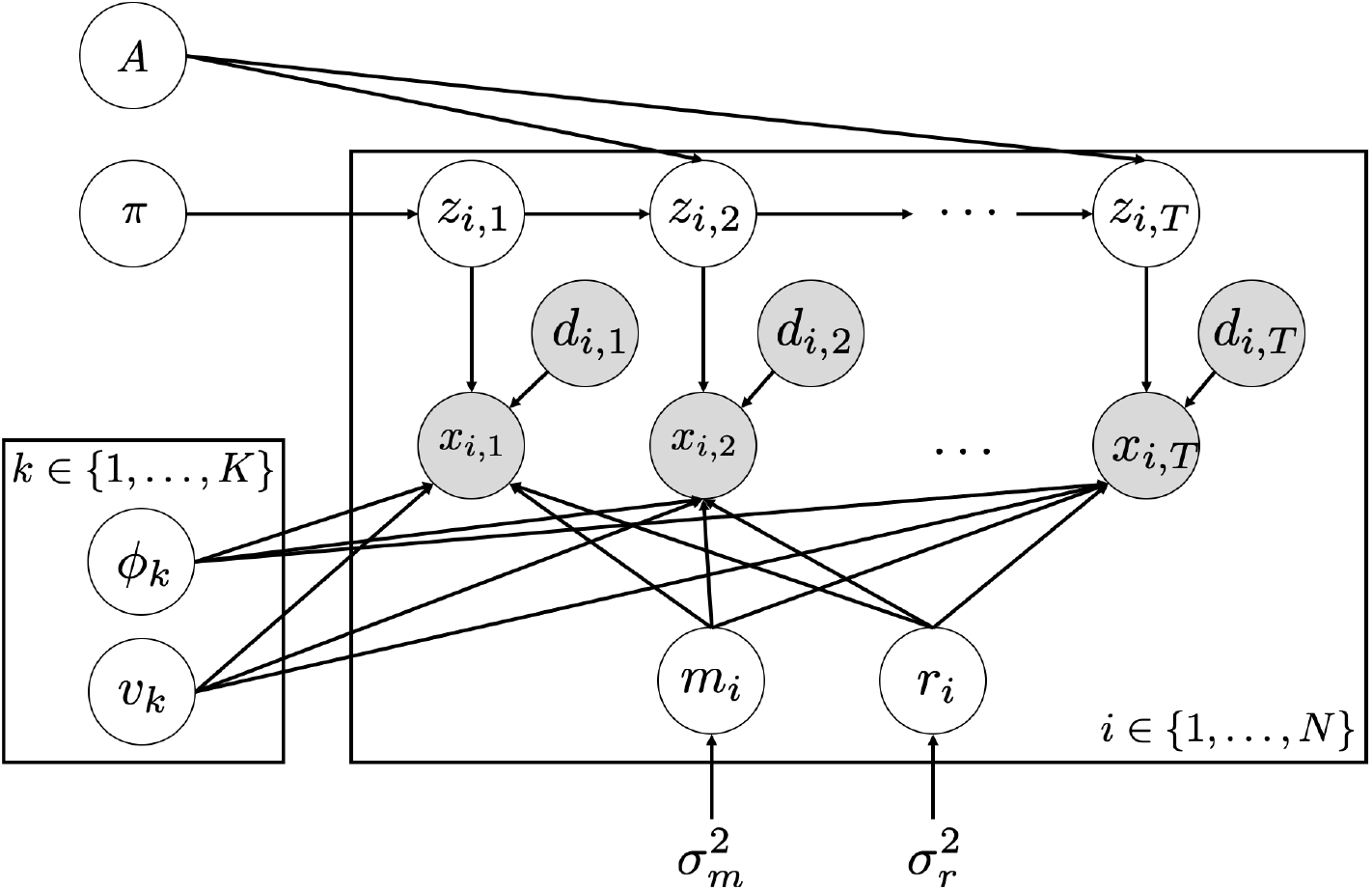
The graph for the proposed model: *z* are the latent states, *x* are the observed variables, *d* are observed medications, *r* are the personalized state effects, and *m* are the personalized medication effects. *ϕ*_*k*_ and *v*_*k*_ are parameters of the observation model. *A* and *π* are parameters of the transition model.

## 3 Learning Algorithm

We use variational methods to learn the proposed model. We approximate the posterior distributions over the *local* latent variables 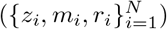 with tractable variational approximations and rely on point estimates for the *global* parameters (*θ*). Our choice of performing point inference for global parameters, those shared by more than one individual, while inferring full distributions for local random variables is motivated by the expectation that local variables which are only informed by a individual’s data would exhibit higher uncertainty. We employ a structured variational approximation that retains the dependence between *z*_*i*_ and *m*_*i*_, *r*_*i*_, and also, crucially, the temporal structure within *z*_*i*_,

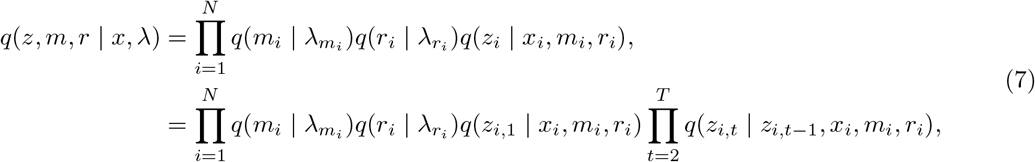

where *λ* are the variational free parameters. We use Gaussians with full covariances to parameterize the variational distributions, 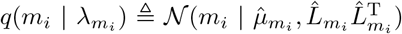 and 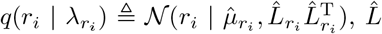. are lower triangular matrice, and use categorical distributions for *z*_*i,t*_.

We minimize the Kullback-Leibler divergence between the variational approximation and the true posterior as well as learn the model parameters, *θ* by maximizing the corresponding evidence lower bound (ELBO),

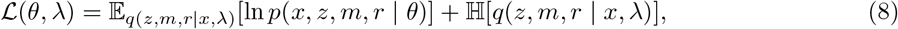

where ℍ [*q*(·)] = − 𝔼_*q*_[ln *q*(·)] is the entropy and the log joint distribution is (see Figure 1),

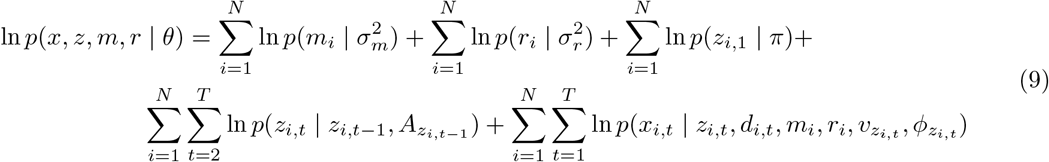

and 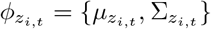.

We maximize the ELBO via coordinate ascent alternating between updates to variational parameters *λ* and model parameters *θ*. Our structured variational approximation, Equation 7, renders expectations required to compute the ELBO intractable. We deal with this issue by using Monte Carlo approximations of the offending expectations and relying on pathwise gradient estimators to differentiate through the sampling process. In particular, we approximate,

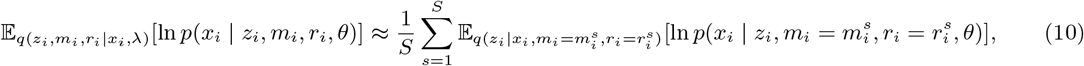

where 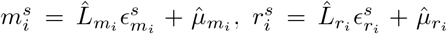 and 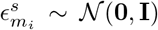 and 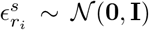. Crucially, we do not need to use a Monte Carlo approximation for evaluating the conditional expectation in Equation 10. Instead we note that we can exactly compute the conditional distribution *q*(*z*_*i*_ | *x*_*i*_, *m*_*i*_, *r*_*i*_), using the forward-backward algorithm [7, 8], a dynamic programming algorithm widely used to learn hidden Markov models, and efficiently compute the conditional expectation by exploiting the Markovian dependencies in the model. These operations are identical to those needed by standard expectation maximization training of HMMs (see appendix for additional details). Given the Monte Carlo expectation we take a stochastic gradient ascent step to update the variational parameters *λ* [9]. Conditioned on *λ*, maximizing the ELBO with respect to *θ* can be done via fixed point updates (see appendix for details). The overall algorithm is summarized in Algorithm 1.

### Algorithm 1 Training procedure of PIOHMM

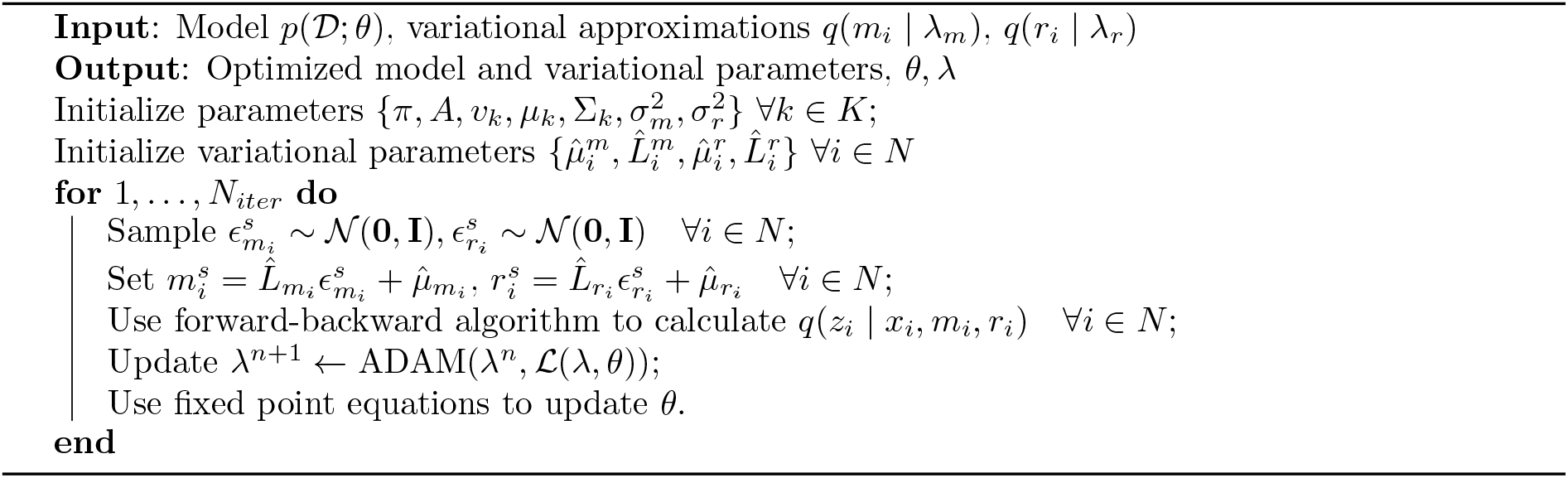

## 4 Related Work

Markov models are not the only class of models that have been considered for disease progression modeling. Several authors have considered Gaussian process (GP) models [3, 10, 11]. GP models are convenient in that they do not require samples to be observed at a fixed rate, are non-parametric probabilistic models, and have been shown to be capable of personalization [10]. However, GP based models are typically unable to discover discrete states of a disease, one of the goals of our work. Similarly, deep learning methods have also been proposed for disease progression modeling [12–14]. While these approaches are able to predict future clinical features, they also do not discover discrete states of progression of a disease. Moreover, they typically need large datasets often requiring tens of thousands of patients records, as is the case in the cited works.

HMMs are well suited to disease progression modeling, particularly when there is an interest in discovering stages of progression. Several studies have leveraged HMMs and variations for disease progression modeling in the past, e.g. [15–17]. Wang et al. [18] proposed continuous time HMMs to model the progression of chronic disease. Sun et al. d[19] leveraged this approach to develop an integrated Huntington’s disease progression model. However none of these previous approaches model system inputs, such as medication, or account for heterogeneity in symptoms between patients in a cohort. Bengio and Frasconi [6] proposed input-output HMMs (IOHMM) as a way to model an observed control signal for a system. In an IOHMM, the inputs influence the observed and/or latent variables. Our work extends IOHMMs to account for medication independent patient heterogeneity while leveraging their ability to model external inputs to account for non disease modifying medications. Altman [20] worked on extending HMM models to account for personalized effects and external inputs. However, unlike us, they relied on Monte Carlo expectation maximization to learn these models, which typically requires running a MCMC sampler within each expectation step and is difficult to scale to both high dimensional data and number of patients. In contrast, our stochastic gradient variational inference approach easily scales along both these dimensions. Finally, recent work by Alaa and van der Schaar [21] proposes to relax the first order Markovian assumption made by typical HMM based disease progression models and personalize progression dynamics across patients. Our contributions are orthogonal, we personalize HMM observation models rather than dynamics. A combination of these two advances comprises an interesting future research direction.

## 5 Results

To demonstrate the algorithm, we apply it to three test cases: a synthetic dataset, a real-world dataset of sensor data from Parkinson’s patients with labeling and a real-world dataset of clinical data from Parkinson’s patients without labeling. In all three cases we demonstrate that the proposed model outperforms other HMM variants.

### 5.1 Synthetic Data

We generate data by considering the following model. First, generate a sequence of latent states governed by the distributions, *z*_*i*,1_ ∼ Cat(*π*), *z*_*i,t*_ | *z*_*i,t−*1_ = *j* ∼ Cat(*A*_*j*_). Then generate observations using, *x*_*i,t*_ | *z*_*i,t*_ = *k* ∼ 𝒩 (*µ*_*k*_, Σ_*k*_). For each sample *i*, a personalized offset is sampled, *r*_*i*_ ∼ Unif[−*b, b*]. Lastly, *structured* noise is added to the observation 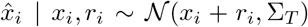, where Σ_*T*_ is specified via a squared exponential kernel,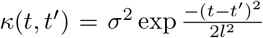. Note that this model is not exactly the same as the proposed model and that when *b* = 0, the observed data is a noisy observation of an HMM with correlated noise.

To simplify the visualization of the results, the dimensionality of the observed data is fixed to one and the number of latent states is two. 100 samples of length 20 are generated as described above and experiments are performed for *b* = 0, 1, 5. The true state means are 0 and 2 and the state variance is 0.1. The results are shown in Figure 2. The experiment demonstrates how the personalized HMM performs no worse than the HMM when personalized effects are not present, and is much better at recovering the state dynamics when personalized effects are present. Because the HMMs is not able to capture personalized differences, the model compensates by inflating the variance of the states and therefore does a poor job of capturing state dynamics.

**Figure 2:**
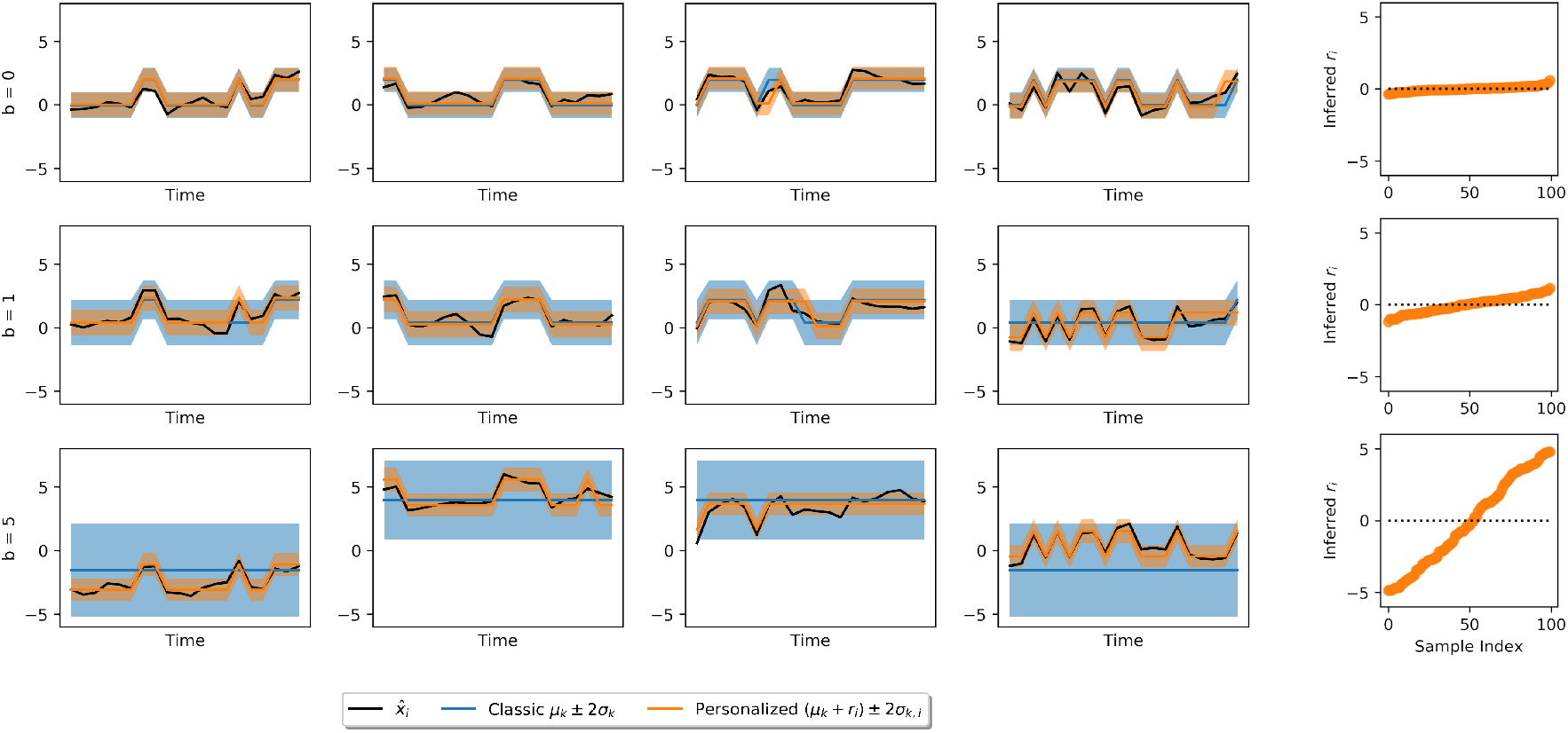
Results for simulated data. The three rows correspond to different distributions of *r*_*i*_ ∼ Unif[−*b, b*] and the four leftmost columns correspond to samples. The rightmost column plots the inferred *r*_*i*_ vs. the sample index. When there are no personalized effects, the personalized and classic HMMs perform approximately the same and the personalized model learns personalized parameters close to zero, as shown in the last column. When a personalized offset does exist, the classic HMM incorrectly assigns states and compensates for the personalization with large variances, as shown in rows two and three. When personalized effects are larger than state effects, a prior to regularize the state effects is appropriate. An analysis of the inferred *r*_*i*_ enables the practitioner to identify samples with large deviations, which may be of special interest.

### 5.2 Application: Parkinson’s Disease

To demonstrate the approach on a real-world dataset, we apply the method to two datasets of Parkinson’s disease patients. As noted in the introduction, Parkinson’s disease (PD) is a chronic neurodegenerative disorder [22]. PD is characterized by a variety of motor and non-motor features and a definitive diagnostic test is not available. Typically diagnosis depends on the presence of specific clinical features, namely rest tremor, rigidity, bradykinesia, and/or postural instability [22, 23]. The heterogeneity of PD symptoms and progression is well-documented but poorly understood [4, 24]. These features make the disease a good fit for our method to learn a clinical representation of disease states as well as how these states evolve in time.

In our first study, we demonstrate the algorithm for the problem of identifying freezing of gait. Freezing of gait is a severe complication of PD resulting in falling, reduced mobility, and increased disability. Identifying objective ways to accurately detect it thus has important clinical implications. In this setting, we have access to ground truth labels from clinical annotators and the relevant states are known: standing, walking, and freezing. In the second study, we apply the algorithm to the more general problem of discovering PD disease states. We describe the resulting states in the context of the current PD literature.

#### 5.2.1 Case Study with Labeled States

##### Data Description

As noted above, the goal of this study was to identify when a patient is experiencing freezing of gait. The study uses the Daphnet Freezing of Gait Dataset, a benchmark dataset which contains information from wearable acceleration sensors [25]. Ten patients are included in the dataset and each has 9 measurements: horizontal forward, horizontal lateral and vertical acceleration from ankle, thigh and trunk sensors. We follow the data processing procedure presented in Bächlin et al. [25] and transform overlapping time windows into the freeze index for each patient and sensor. Two patients have no freezing of gait events and are removed from our study. The time trajectory of each patient is then divided into training and testing sequentially with the first 400 time steps (200 seconds) used for training and the subsequent 400 time steps used for testing.

##### Model Description

For this application, we choose the observation model

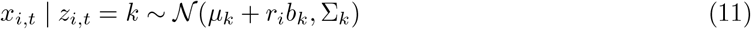

which has personalized state effects. This is motivated by the original study which notes that their detection model has variable performance due to the different walking styles of the patient. There are three pre-defined states: standing, walking, and freezing of gait. We do not expect personalized differences in standing, therefore we introduce a variable *b* = [0, 1, 1], such that *r*_*i*_ has no effect for one of the states. Note that beyond specifying *b*, no special care is taken in initializing the model during training. A second model using a classic hidden Markov model is also trained for comparison. This model has the observation model

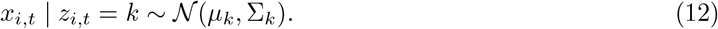

##### Results

The two models are compared using several metrics. The test log likelihoods are -238181 and -238660 for the personalized and standard models, respectively, implying the personalized model is better at describing the data. Figure 3 shows an example of the freeze index for one sensor and the corresponding prediction for patient 9 along with model metrics. The receiver operating curve is calculated for each model using the belief state of the freezing state

**Figure 3:**
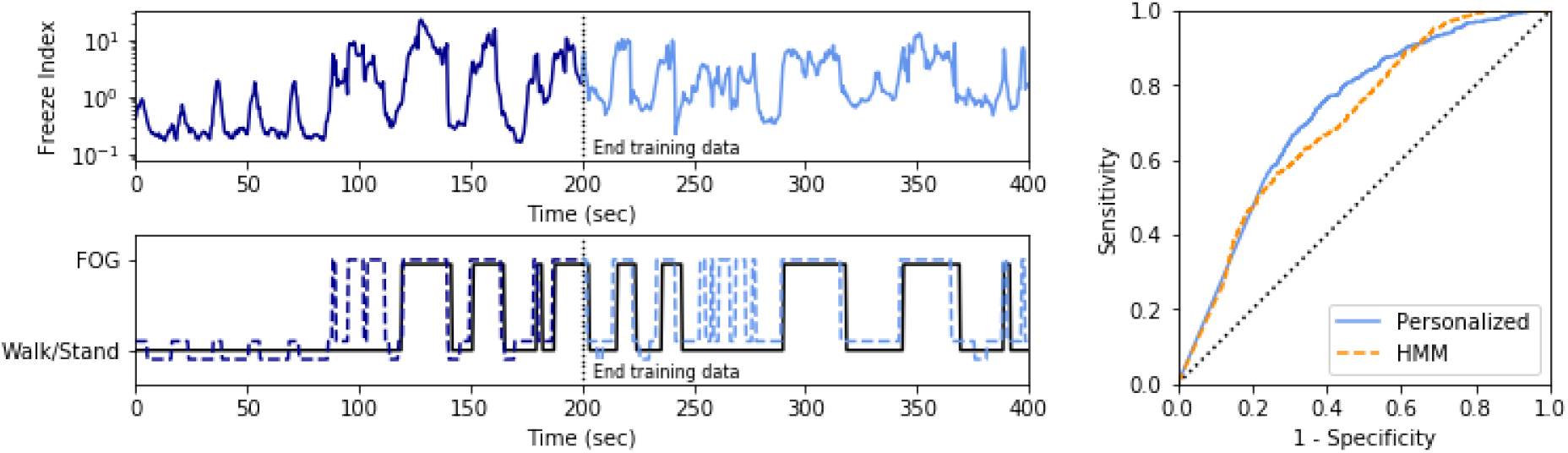
Results from the case study with labeled states. Moving from left to right, the upper plot shows the freeze index for the ankle sensor acceleration in the vertical direction. The lower plot shows the state prediction (dotted line) compared to ground truth (solid line). Note that the ground truth labels do not differentiate different tasks and only have information for freezing of gait (FOG) or no freezing of gait. The right plot shows the receiver operator curve for the test data using the personalized and standard hidden Markov models using the belief state for the freezing state. The AUCs are 0.725 and 0.705, respectively.

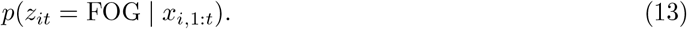

The AUCs are 0.725 and 0.705 for the personalized and standard model, respectively, indicating slightly better performance for the personalized model.

Qualitatively, the inferred personalized state effects, *r*_*i*_, can be used to identify outlier patients, who may be of interest. Here, the largest *r*_*i*_ corresponds to the patient with the largest Hoehn and Yahr score, a measure of PD severity not used in model training.

#### 5.2.2 Case Study without Labeled States

##### Data Description

For this study, we use the Parkinson Progression Marker Initiative (PPMI) dataset [26]. PPMI is an observational, longitudinal, multi-center study that enrolled 423 PD patients. PPMI collects clinical, imaging, and biospecimen samples. In this analysis, we focus on the clinical assessments, specifically those measured via the Movement Disorder Society Unified Parkinson’s Disease Rating Scale (MDS-UPDRS) [27]. The MDS-UPDRS is a four part assessment that contains a combination of patient reported- and physician-assessed measures. The four parts are: (I) non-motor experiences of daily living, (II) motor experiences of daily living, (III) motor examination, and (IV) motor complications. Each item on the scale is rated from 0 (normal) to 4 (severe). For our study, we do not use part IV therefore the observed data has 59 dimensions.

Levodopa is the primary medication used to treat Parkinson’s disease and is thought to have no disease-modifying effect [28]. Levodopa is a dopaminergic medication and is primarily used to treat motor symptoms. Levodopa may be administered in concert with other medications such as a dopamine agonist, monoamine oxidase type B inhibitor, or a catechol-O-methyl transferase inhibitor. In order to model the combination of these drugs in a consistent manner, the levodopa equivalent daily dose (LEDD) [29] has been developed. LEDD information is provided in PPMI and used to model medication effects in our study. Per the PPMI protocol, PD patients must not have started medications at time of enrollment and initiate different medications at varying times over the study, depending on specific patient characteristics. The dose as well as the response changes over time. Medication effects are captured to some degree by motor examination occurring prior to intake of levodopa or dopamine agonists, and repeated again 1-4 hours after medication are taken. This process is referred to as ‘on-off’ testing.

##### Model Description

Based on domain understanding of Parkinson’s disease, we choose the observation model

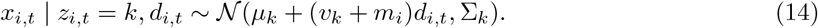

This model captures our prior beliefs that there are several unknown disease states whose observations are impacted by medication use. We expect that the impact of medication is a function of both the individual and the disease state. The transition matrix, *A* is constrained to be upper triangular to enforce progressive disease states.

A full distribution of the number of visits per patient is available in the appendix; on average, the patients have observations through 21 visits and a total of 12 observed visits. This discrepancy is a result of the PPMI protocol, which increases the time between visits as the time since enrollment increases. Because the protocol establishes a visit schedule prior to patient enrollment, a missing at random assumption is reasonable. We note that it is possible that patients deviate from their schedule based on their disease status but did not find evidence to that effect and proceeded with an assumption of data that is missing at random (see appendix for more detail). The time step is fixed to three months and any missing values are marginalized. The dataset is divided into training and testing with 333 patients used for training and 83 patients used for testing. Patients with only one observation were excluded (N=7). To compare with the proposed PIOHMM, standard HMM and IOHMM models are also developed. For all models, we use a 5-fold cross validation strategy on the training data to select the number of states.

##### Results

Figure 4 plots the test log-likelihood per patient for the three models. The PIOHMM outperforms the other models, suggesting that the PIOHMM is a better model for the data.

**Figure 4:**
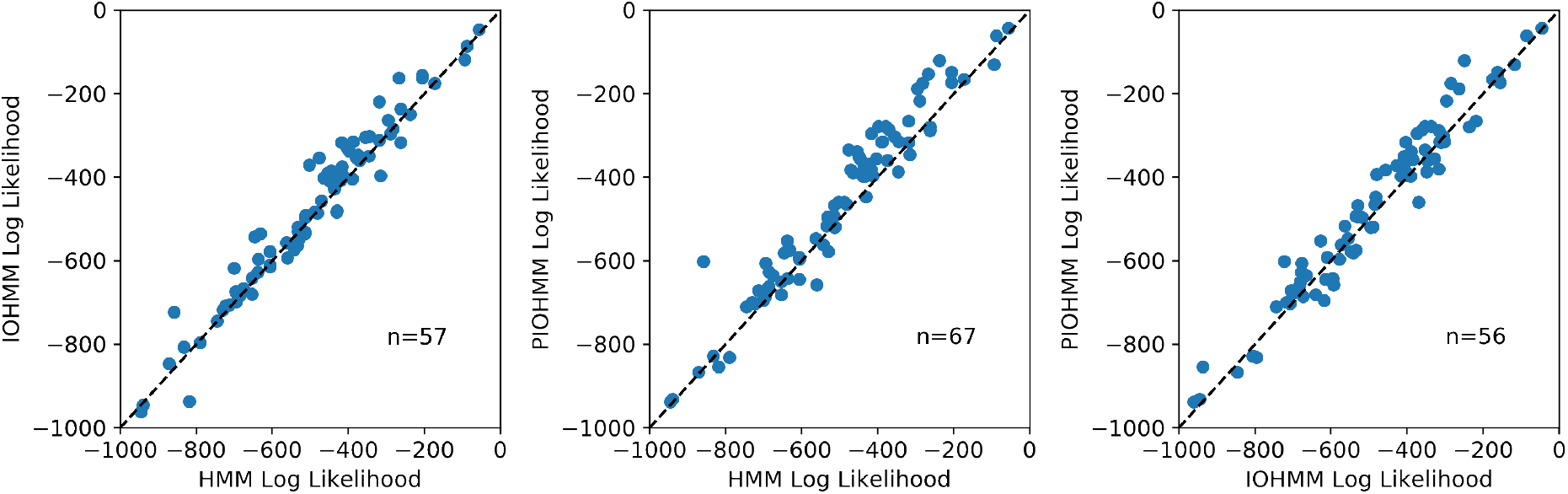
Scatter plots showing the per sample test log-likelihood (N=83) for the three different Markov models. The number on the plot indicates the number of samples that are above the diagonal line. These results show that the PIOHMM improves upon both the IOHMM and HMM in terms of test log likelihood.

By modeling personalized-medication and state-medication effects, we hope to recover relevant clinical latent states. To characterize the states, we perform a post-hoc analysis, focusing on the primary clinical symptoms used for diagnosis: tremor, bradykinesia, rigidity and postural instability/gait (PI/G). See Figure 5 for a summary. Note that the states are ordered in terms of progression, e.g. state 1 cannot transition to state 0, state 2 cannot transition to states 0 or 1, etc. State 1 is the most frequent state at enrollment and has the lowest total MDS-UPDRS score. State 0 has no patients on medication. States 2 and 4 both have moderate tremor and are primarily differentiated by which side of the body the disease affects. State 6 has high tremor. States 3, 5, and 7 have increasingly severe gait issues. Per the subtype methodology of Stebbins et al. [30], which uses MDS-UPDRS subitems to assign patients who are not on medication to tremor or PI/G subtypes, state 5 is PI/G dominant, states 0-4 and 6 are tremor dominant, and state 7 is indeterminate. Unlike these definitions, the learned disease states have more granularity and capture severity.

**Figure 5:**
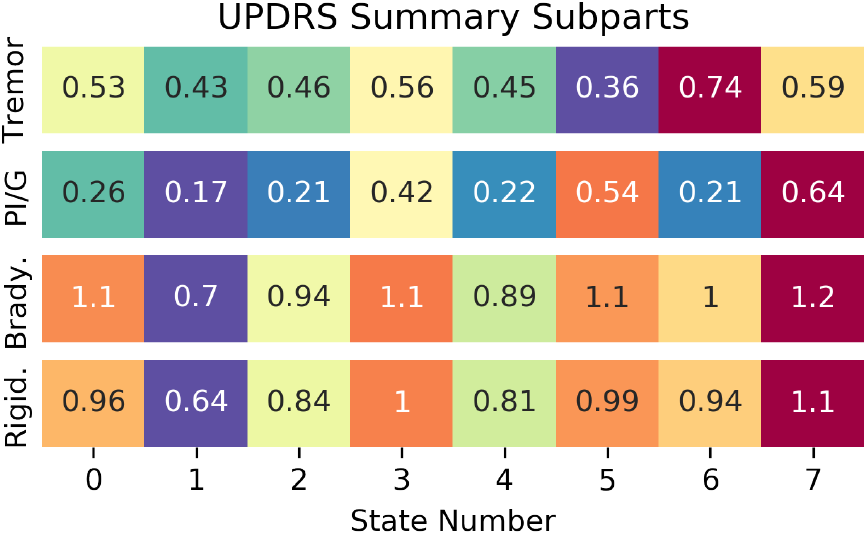
Average of the subitems corresponding to primary PD symptoms from the state means, *µ*_*k*_. The subscores are calculated using the MDS-UPDRS as follows: tremor, 2.10, 3.15-3.18, postural instability gait, 2.12-2.13, 3.10-3.12, bradykinesia, 3.4-3.8, 3.14, and rigidity, 3.3. The colors correspond to the range of severity within each category (blue for the least severe and red for the most severe) and are redundant with the value in the heatmap.

In addition to this static view of the learned states, we are also interested in how these states evolve in time. Analysis of the transition probabilities, *A*, provides insight to expected transition patterns. Using the Viterbi algorithm, marginalizing over the personalized effects, we can also estimate the most probable sequence for each patient. As a demonstration of how the model could be used, we predict whether a patient in the test dataset will be in a tremor dominant or postural instability/gait dominant subtype based on the definitions of Stebbins et al. [30]. To do this, we use the state assignment probabilities at study entry and the transition matrix to calculate state assignment in one year. The probabilities at one year are then used to calculate a weighted average of MDS-UPDRS subitems. Unlike the states from our model, these literature subtypes are only defined for patients not on medications, therefore only 24 of the 83 patients in the test set have the required information. Using the model, 17 patients are correctly predicted. If instead we assume that patients do not change subtypes from study entry, only 14 patients are correctly predicted. This heterogeneity is supported by independent analysis of the PPMI data [5].

We can also analyze the parameters that interact with medication to gain new insights. Overall, facial expression, consistency of rest tremor, and body bradykinesia have the most positive response (i.e. largest decrease) as a function of LEDD, turning in bed and sleep (sleep problems and daytime sleepiness) have the most negative response (i.e. largest increase), and freezing has the smallest response (i.e. approximately no change). These results are independently verified by ‘on-off’ paired testing that occurs elsewhere in the PPMI dataset, data not used in the model. In that dataset, consistency of rest tremor had the most positive response and freezing of gait was the least responsive (note that sleep issues are not measured as part of the test). Issues regarding sleep have been documented as a possible side effect of levodopa in PD in the past [31, 32]. An analysis of the personalized effects shows that the change in score for turning in bed has the greatest variability. The relationship between turning in bed and levodopa has previously been studied and is thought to be responsive to levodopa therapy but not always consistently [33]. It is interesting to note that this measurement in the MDS-UPDRS is self-reported. These insights lead to interesting hypotheses to support further investigation which would not have been otherwise revealed.

## 6 Discussion

Disease progression models are important computational tools in healthcare, however, they face particular challenges as compared to general time series models. To address these challenges, we propose a probabilistic model which is capable of accounting for personalized state effects, personalized medication effects, state-based medication effects, or any combination thereof while learning disease states. We refer to this model as a personalized input-output hidden Markov model (PIOHMM). We demonstrate the model’s success on synthetic and real-world datasets. The model is well-suited to the scenario where a disease progresses heterogeneously and a characterization of disease states is a modeling goal.

### Limitations

While the proposed model is a reasonable first step several of its assumptions could be relaxed. First, the model as presented does not account for the effect of demographic factors on the progression of the disease. When such information is available, one way of incorporating it in the model would be to define the personalized effects to be a function of the demographics, *r*_*i*_ = *w*^*T*^ *y*_*i*_, where *y*_*i*_ are the available demographics for patient *i*. Next, we have chosen to model medications as only impacting the observation model and not the transition model. This is motivated by the large number of complex diseases which do not have disease-modifying therapeutics but do have medications to alleviate symptoms in patients. However, there may be scenarios where such an assumption is inappropriate, for instance, when a disease modifying drug is available. We could adapt the model to account for transition models that are a function of medication or other covariates, for instance *z*_*i,t*_ | *z*_*i,t−*1_, *d*_*i,t−*1_ ∼ Cat(*𝒮* (*A*_*j*_*d*_*i,t−*1_)), where *𝒮* (·) represents a softmax transformation. Such a medication or patient specific covariate conditioned transition model would also relax the assumption that the disease dynamics, as specified by the HMM transition matrix, is shared among all patients. Finally, it is possible that the medication effects are not linear and that medication does not affect all disease measures but only a subset which needs to be discovered from the data. In such situations, one could model non-linearity through a non-linear basis function expansion and use an automatic relevance detection [34] or other sparsity inducing priors to infer the active medication effects.

## Data Availability

Data used in this study is open source and available via ppmi-info.org and http://archive.ics.uci.edu/ml/datasets/Daphnet+Freezing+of+Gait

## 7 Acknowledgments

The authors would like to thank Mark Frasier for helpful comments and discussion. Data used in the preparation of this article were obtained from the Parkinson’s Progression Markers Initiative (PPMI) database (www.ppmi-info.org/data). For up-to-date information on the study, visit www.ppmi-info.org. PPMI – a public-private partnership – is funded by the Michael J. Fox Foundation for Parkinson’s Research and funding partners, including abbvie, Allergan, amathus therapeutics, Avid radiopharmaceuticals, Biogen, BioLegend, Bristol-Myers Squibb, Celgene, Denali, GE Healthcare, Genentech, GlaxoSmithKline, Golub Capital, Handl Therapeutics, insitro, Janssen, Lilly, Lundbeck, Merck, Meso Scale Discovery, Pfizer, Primal, Prevail Therapeutics, Roche, Sanofi Genzyme, Servier, Takeda, Teva, UCB, Verily and Voyager Therapeutics.

## A Forward-backward algorithm details

The application of the forward-backward algorithm follows the steps of the forward-backward algorithm as applied to an HMM. We define

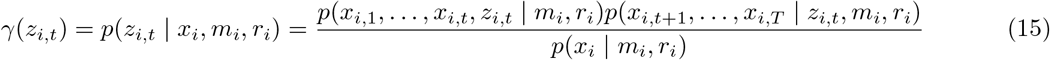

Further, we define

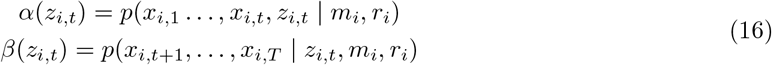

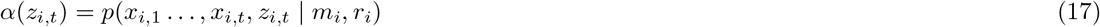

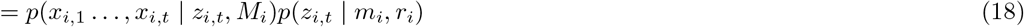

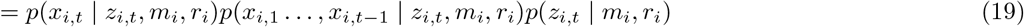

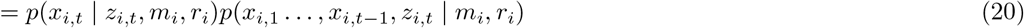

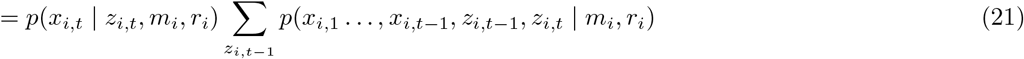

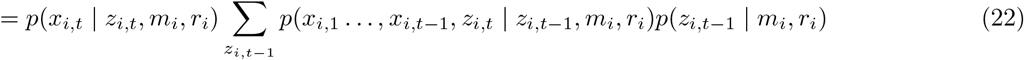

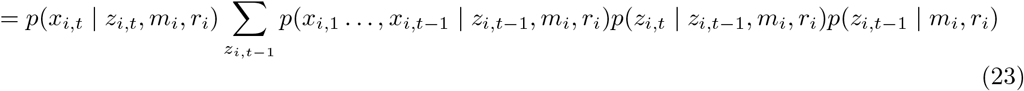

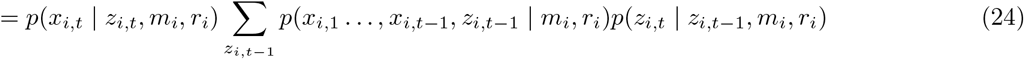

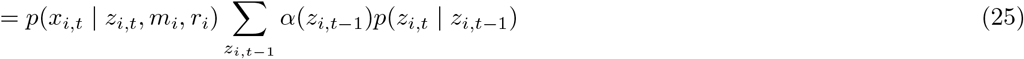

Similarly,

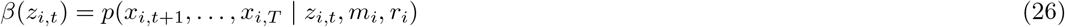

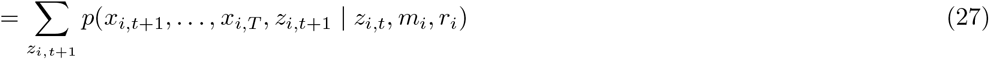

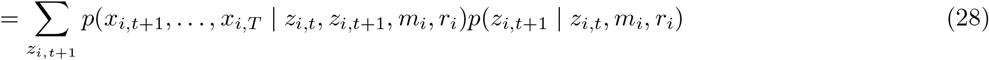

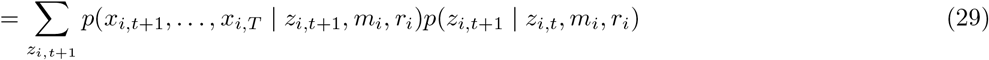

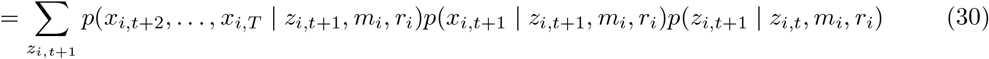

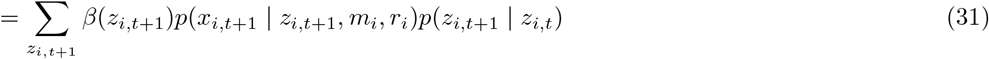

The recursion is started using

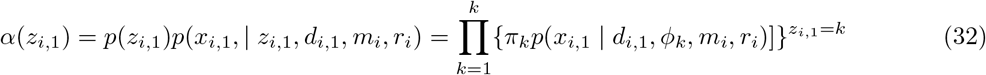

and

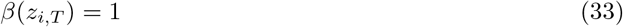

The objective function can be expanded and written

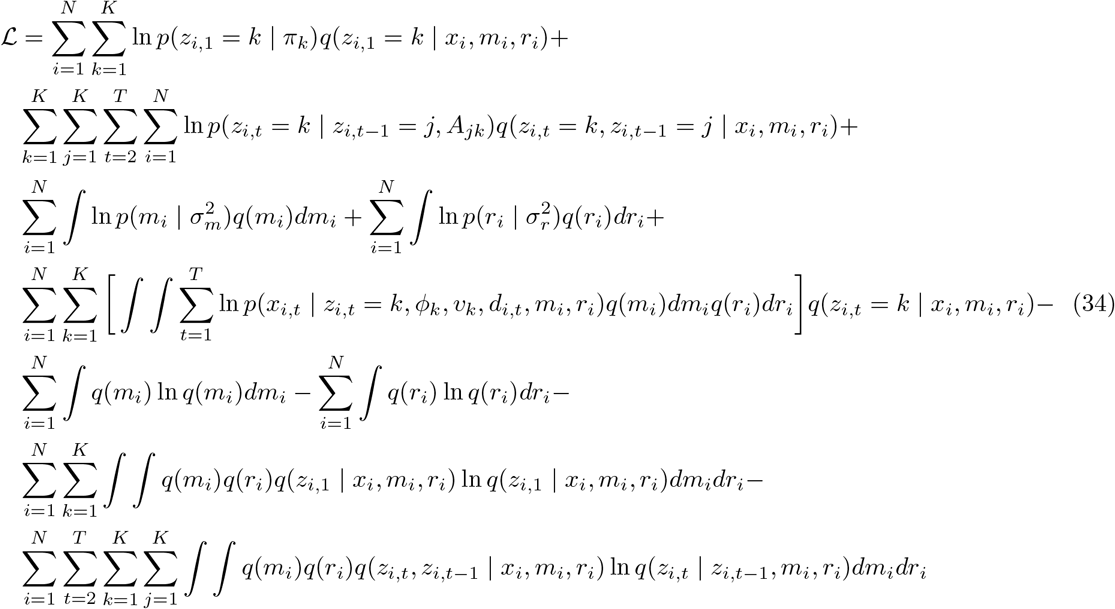

## B Fixed point updates

The equations for the parameter updates under the observation model

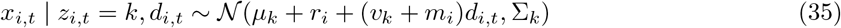

are shown below.

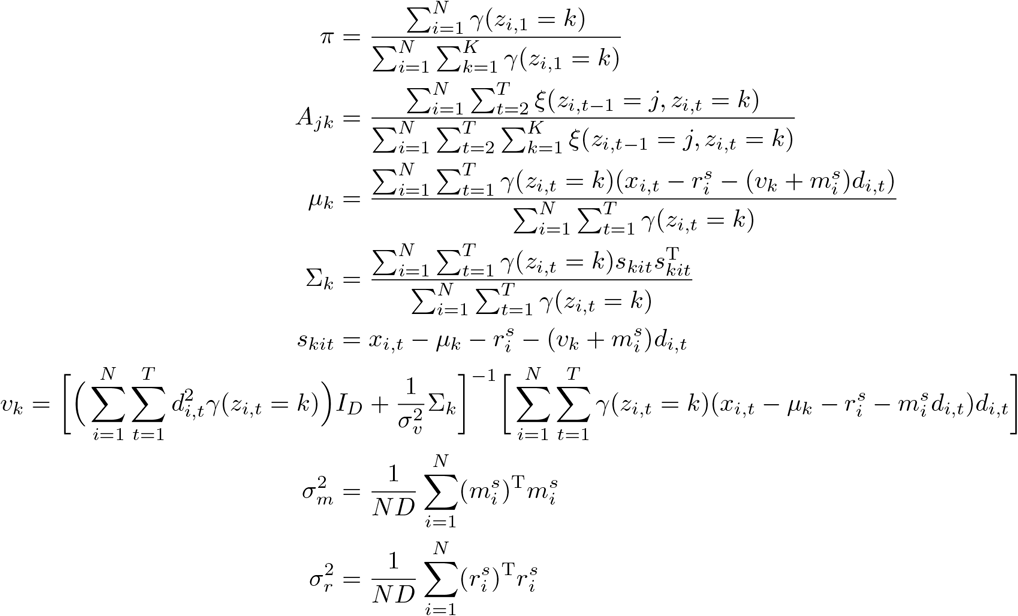

## C Parkinson’s disease patient cohort

### C.1 Data availability

Figure 6 shows the distribution of the number of visits per patient. On average, data is observed up to visit 21, although each visit within the trajectory is not necessarily observed. The average number of observations per patient is 12.

**Figure 6:**
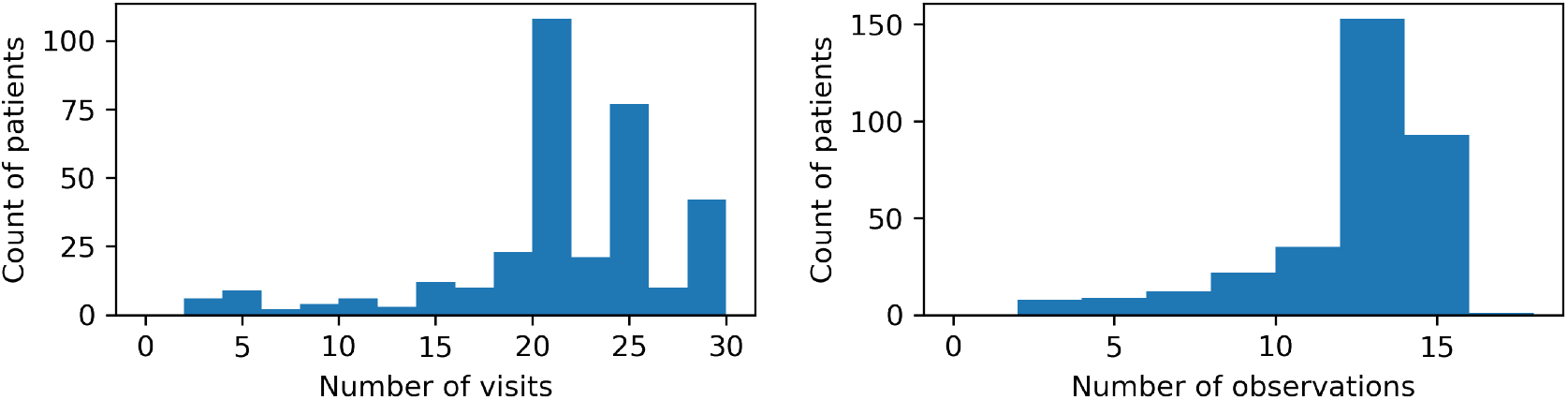
Distributions of the index of the last observed visit and the number of observations per patient. Not all visits for a trajectory may be observed. The left histogram represents the time point of the final observation for each patient in the training set and the right histogram represents the number of observed visits. This is a result of the observation protocol. The testing data is distributed similarly.

### C.2 Missing data analysis

Although there is no robust test to confirm that data is not missing at random, empirical evidence suggests this is not the case for the PPMI PD dataset. We show the trends in number of total observations as a function of time since study enrollment as well as MDS-UPDRS total and change in MDS-UPDRS total in Figure 7. If data were not missing at random, we might hypothesize that patients who have high MDS-UPDRS total values at last visit or patients who have large increases in MDS-UPDRS would have a small number of visits due to study drop-out. For the most part, we do not observe any linear trend (*r*^2^ = 0.01 for total and *r*^2^ = 0 for change), whereas there is a strong linear trend in the relationship between time since enrollment and number of visits (*r*^2^ = 0.83). Although inconclusive, we take this to imply that a majority of missingness is due to the study protocol and not censoring or drop-out as a function of disease state.

**Figure 7:**
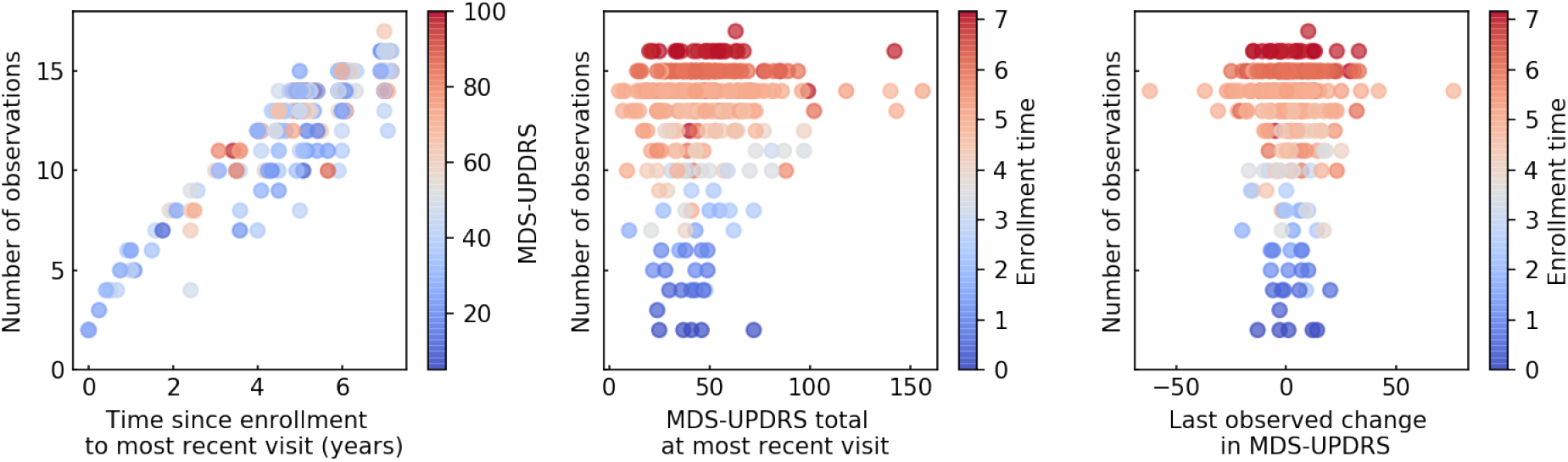
Trends of total number of observations with various patient attributes. Time since enrollment is strongly correlated with the total number of observations whereas measures of disease state are not.

### Footnotes

1 To arrive at IOHMM from PIOHMM, set *r*_*i*_ = 0, and *m*_*i*_ = 0.

